# Predicted failure of common Mpox PCR testing on a recent DRC clade I variant: Persistent genomic surveillance is critically important for accurate diagnostics

**DOI:** 10.1101/2024.07.22.24310820

**Authors:** Dawn Gratalo, Valerie J. Morley, Ben Oppenheim, Casandra W. Philipson, Sarah Rush, Swati Sureka, Mitchell I. Wolfe, Birgitte B. Simen

## Abstract

Pathogens can rapidly evolve to evade detection via existing diagnostic testing capabilities, creating blind spots for health care and security systems. Persistent genomic surveillance of pathogens can help identify and address these gaps. Here, we present a case study of a new lineage of Monkeypox virus Clade I that emerged during an outbreak in the Democratic Republic of the Congo, in which a genomic deletion allowed the virus to evade detection by current diagnostic assays. We rapidly developed and validated an updated assay. Availability of pathogen genomic data is a critical input, and often a bottleneck, to response efforts.

## Background: Effective pathogen diagnostic tools depend on genomic surveillance

Traditional pathogen testing relies on PCR-specific primers for clade identification. These tests use either single or multi-primer designs depending on the manufacturing of the diagnostic. Although multiple PCR targets provide redundancy in testing, implementing this approach is not always feasible due to resource constraints or variability within the genome. A tradeoff is often made by limiting the design to a single primer pair. This creates a potential deficit in detection, as a pathogen can go undetected if its genetic composition mutates, which in turn can lead to extended periods of undetected pathogen transmission. This problem may be magnified in low-resource settings, due to gaps in surveillance systems and reduced access to health services.

Single primer tests are especially problematic since mutations in the genome can lead to complete test failure. During the COVID-19 pandemic, mutations in the SARS-CoV-2 spike gene (S-gene) caused failure or reduced sensitivity in widely used commercial kits, depending on the number of primer sets and the associated interpretation rules. It was eventually recognized that S-gene dropout or S-Gene Target Failure (SGTF) created a unique signature that could be used to identify shifts in circulating variants including the introduction of the omicron saltation variant. During this time, the scientific community engaged in an unprecedented degree of open sharing around protocols and findings, which allowed such discoveries to be made rather quickly.

There are additional examples of pathogens that have escaped detection, resulting in increased transmission, failed diagnostic testing, and resultant delays in mitigation. A variant of *Chlamydia trachomatis* circulated undetected in Sweden for 3 years after widely used molecular tests from multiple manufacturers failed to detect the new variant.^1^ Only after additional genetic characterization was a particular 377-bp deletion discovered as the source of test failure. In another example, of malaria testing in Africa, it was discovered that mutations in the *Plasmodium falciparium* histidine-rich protein 2 or 3 (pfhrp2/3) gene allowed strains to escape detection, resulting in a ∼10% false negative rate in diagnostic testing. Genomic sequencing identified mutational patterns and showed evidence of evolutionary pressure favoring pfhrp2-deletions, underscoring the importance of genomic sequencing for variant detection and transmission and virulence tracking.^2,3^

To avoid potential gaps in pathogen detection and enable rapid and effective outbreak mitigation, robust genomic sequencing is needed. Sequencing provides essential information that can identify mutations that may affect PCR-based testing, allowing diagnostics to be updated with new designs as variants emerge. Below, we present a recent instance in which sequencing data of a novel Mpox strain revealed a critical gap in existing diagnostic testing, enabling us to rapidly develop an updated assay.

### Case study: A novel lineage of Monkeypox virus Clade I in Kamituga, DRC

Monkeypox virus (MPXV), the causative agent of Mpox disease, is an enveloped double-stranded DNA virus belonging to the *Orthopoxvirus* genus, with two genetic clades, Clade I (Congo Basin) and Clade II (Western Africa). Prior to the 2022 global epidemic caused by Clade II, the majority of reported Mpox cases were caused by MPXV Clade I, with the Democratic Republic of the Congo (DRC) having the highest burden of disease among affected countries.^4^

MPXV Clade I is of significant concern due to its high case fatality rate – estimated at up to 10%, as compared to <1% for MPXV Clade II.^5,6^

Clade I remains a significant public health concern in DRC, and a national outbreak has been ongoing since December 2022. In April 2023, the World Health Organization (WHO) reported the first known instances of Clade I being associated with sexual transmission. Since the start of 2024, 7,851 cases have been reported in DRC with an estimated case fatality rate of 4.9%^7^.

In April 2024, a new lineage of clade I MPXV with epidemic potential was reported based on twenty-two viral genome sequences from an October 2023 outbreak in Kamituga health zone, within DRC’s South Kivu province. The Kamituga MPXV outbreak spread rapidly, with 241 cases reported in 5 months.^8^ It was further reported that the Kamituga sub-lineage contained a 1kb deletion in the viral genome that allows the virus to evade detection by current diagnostic tests, resulting in a critical gap in MPXV surveillance capabilities.^8,9^

Full genome alignment of the MPXV Clade I reference (Genbank NC_003310.1) and Kamituga sub-lineage allowed us to rapidly develop a PCR primer redesign that captures both the reference genome and the Kamituga sub-lineage (Figure 1), highlighting the criticality of genomic sequencing data. The updated assay was validated and available for deployment within three weeks of sequence reporting. (Figure 2)

**Figure 1:**
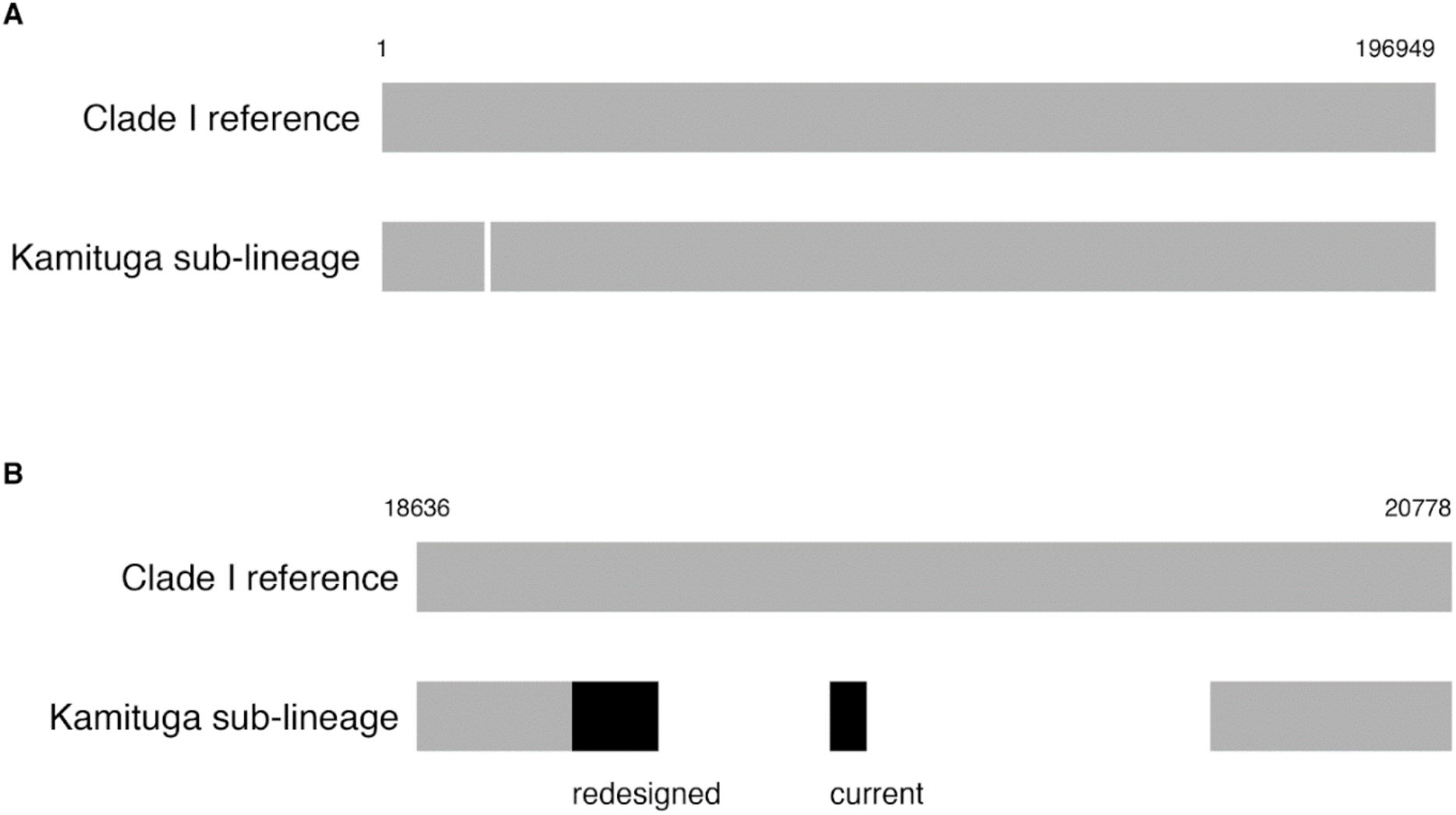
Genomic map of MPXV PCR assay designs. A) Full genome alignment of MPXV Clade I reference (Genbank NC_003310.1) and Kamituga sub-lineage. The Kamituga sub-lineage has a deletion of ∼1kb relative to the reference (19136-20278). B) PCR amplicons (black) mapped against relevant MPXV genomic region. The current amplicon falls within the Kamituga deletion, and a redesigned primer captures both the reference genome and the Kamituga sub-lineage.

**Figure 2.**
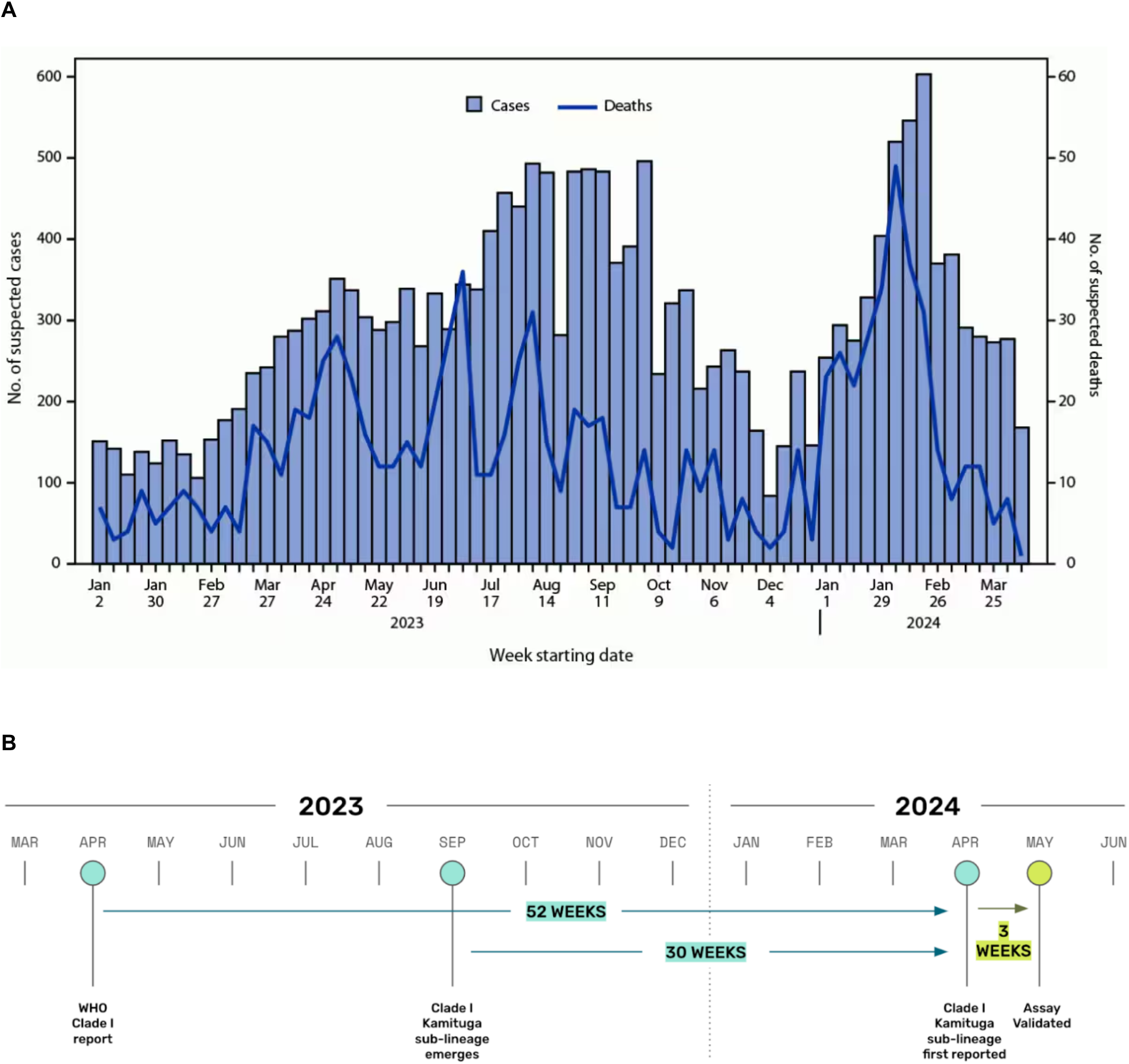
A) Suspected clade I mpox cases and deaths* — Democratic Republic of the Congo, January 1, 2023–April 14, 2024 McQuiston JH, Luce R, Kazadi DM, et al. U.S. Preparedness and Response to Increasing Clade I Mpox Cases in the Democratic Republic of the Congo — United States, 2024. MMWR Morb Mortal Wkly Rep 2024;73:435–440. DOI: http://dx.doi.org/10.15585/mmwr.mm7319a3 B) timeline of outbreak 2023-2024

## Methods

The PCR detection assay design was based on a multiple sequence alignment created from publicly available sequences using MAFFT v7.520. The alignment included Clade I sequences from the DRC outbreak^8^ with and without the ∼1kb deletion and all Clade I sequences available in NCBI Genbank. The primer design was examined for cross-reactivity by BLASTn analysis against standard databases (NCBI default settings) and, to ensure Clade I specificity, further aligning primer sequences against representative Clade II genomes (NC_063383.1, MN346691, OL504742.1, OP535326.1) and related orthopoxviruses (NC_006998.1, NC_001611.1, NC_003663.2). No significant homology to other targets was found. Primer and probe concentrations were optimized for dPCR testing on the QIAcuity Digital PCR System (QIAGEN) with multiplex compatibility evaluated in a panel that included standard Clade I and Clade II assays.

## Discussion: Lessons for public health surveillance systems

Persistent genomic surveillance is a necessary complement to diagnostic testing to keep pace with pathogen evolution. Effective diagnostic capabilities are especially critical during the emergence of a novel strain with epidemic potential in order to monitor and contain the outbreak. As this MPXV case study demonstrates, pathogen genomic analysis can be a rate-limiting step to developing and updating diagnostics during this consequential period. Public health policymakers and responders must focus on deploying biosurveillance networks that include robust and timely genomic next generation sequencing.

In addition to providing early warning variant detection, genomic sequencing can also be used to identify characteristics that may lead to increased pathogenicity, symptom severity, and therapeutic resistance.^10,11,12^ This is especially important in regards to MPXV vaccine design, where the pathogen has very high morbidity and mortality potential, and sequencing data can inform vaccine updates or give early warnings of escape variants.^13,14^

Lastly, it is particularly important to intensify data sharing for pathogens with poorly understood genetic epidemiology and known potential for mutation that can lead to more frequent disease spillover or increased human-to-human transmission. We urge the public health community to strengthen policies and mechanisms for improved testing, data sharing, and genomic surveillance efforts in the monitoring of this and all emerging pathogens.

## Data Availability

All data produced in the present study are available upon reasonable request to the authors

